# Prediction of in-hospital mortality among intensive care unit patients using modified daily Laboratory-based Acute Physiology Scores, version 2 (LAPS2)

**DOI:** 10.1101/2023.01.19.23284796

**Authors:** Rachel Kohn, Gary E. Weissman, Wei Wang, Nicholas E. Ingraham, Stefania Scott, Brian Bayes, George L. Anesi, Scott D. Halpern, Patricia Kipnis, Vincent X. Liu, R. Adams Dudley, Meeta Prasad Kerlin

## Abstract

**Background:** Mortality prediction for intensive care unit (ICU) patients frequently relies on single acuity measures based on ICU admission physiology without accounting for subsequent clinical changes.

**Objectives:** Evaluate novel models incorporating modified admission and daily, time-updating Laboratory-based Acute Physiology Scores, version 2 (LAPS2) to predict in-hospital mortality among ICU patients.

**Research design:** Retrospective cohort study.

**Subjects:** All ICU patients in five hospitals from October 2017 through September 2019.

**Measures:** We used logistic regression, penalized logistic regression, and random forest models to predict in-hospital mortality within 30 days of ICU admission using admission LAPS2 alone in patient-level and patient-day-level models, or admission and daily LAPS2 at the patient-day level. Multivariable models included patient and admission characteristics. We performed internal-external validation using four hospitals for training and the fifth for validation, repeating analyses for each hospital as the validation set. We assessed performance using scaled Brier scores (SBS), c-statistics, and calibration plots.

**Results:** The cohort included 13,993 patients and 120,101 ICU days. The patient-level model including the modified admission LAPS2 without daily LAPS2 had an SBS of 0.175 (95% CI 0.148-0.201) and c-statistic of 0.824 (95% CI 0.808-0.840). Patient-day-level models including daily LAPS2 consistently outperformed models with modified admission LAPS2 alone. Among patients with <50% predicted mortality, daily models were better calibrated than models with modified admission LAPS2 alone.

**Conclusions:** Models incorporating daily, time-updating LAPS2 to predict mortality among an ICU population perform as well or better than models incorporating modified admission LAPS2 alone.

## INTRODUCTION

Time-updating acuity assessment is critical for clinical decision-making, quality and safety initiatives, hospital performance, and clinical research. Multiple mortality prediction scores incorporating acute physiologic data have been developed and validated among intensive care unit (ICU) patients. However, these scores do not capture changes in acuity over time, which may add predictive value, and those that do tend to have narrow ranges, potentially limiting discrimination.^1–5^

Escobar et al. developed the Laboratory-based Acute Physiology Score, version 2 (LAPS2), a score assigned at hospital admission ranging from 0-414 to predict mortality among hospitalized adults.^6,7^ LAPS2 uniquely includes administrative and physiologic variables from the 72 hours preceding admission, data readily available within the integrated health system in which it was developed. Additionally, LAPS2 utilizes a preliminary model to subdivide the population into low and high mortality risk in order to impute missing data accordingly, rather than assuming normal values as is common in other mortality prediction scores.^7^

LAPS2 performs better than many other mortality prediction models. Furthermore, recent work comparing LAPS2 to the commonly used Sequential Organ Failure Assessment (SOFA) score demonstrated that LAPS2 is more racially equitabe,^8^ making it an exemplar model for further expansion and refinement. However, several features limit its utility in ICU populations. First, most hospitals lack integration with outpatient sites to capture the required data from 72 hours prior to hospital presentation. Second, admission acuity does not account for informative changes in patients’ physiology. Finally, LAPS2 has not been comprehensively studied among ICU patients, for whom mortality risk is highest and updated prognostication is most critical to decision-making. Therefore, our objective in this study was to develop and compare novel prediction models using daily LAPS2 based exclusively on data collected after hospital presentation to predict in-hospital mortality among ICU patients.

## METHODS

### Study overview, sites, and population

We created a retrospective cohort of patients ≥18 years old admitted to an ICU of four hospitals of one health system between October 1, 2017, and September 30, 2019; and a fifth hospital between June 1, 2018, and September 30, 2019 (when that hospital integrated into the health system). The five hospitals vary in size (approximately 250 to 700 beds), location (two suburban and three urban), and academic affiliation (two primary sites for multiple large residency programs, one primary site for a few small residency programs, and two without trainees). The study was approved by the University of Pennsylvania Institutional Review Board.

### Specification of admission and daily LAPS2 scores

For each patient, we estimated a modified LAPS2 for hospital admission adapted from the original specification to include data obtained exclusively after hospital admission (**Supplement**). To align with shift schedules, we defined hospital and ICU days as 7:00am to 6:59am the following calendar day. We estimated admission LAPS2 in two steps. First, we built a logistic regression model (pre-LAPS2 model) including patient age, gender, hospital admission source, and values at the end of the first hospital day of serum sodium, blood urea nitrogen (BUN) to creatinine ratio, and anion gap to serum bicarbonate ratio to predict in-hospital mortality, stratifying patients into high (≥6%) or low (<6%) mortality risk. Second, we used the worst values of all LAPS2 variables from the first hospital day to assign points as in the original LAPS2 specification, imputing a normal value for low mortality risk patients and an abnormal value for high mortality risk patients when values were missing (**Supplement**).^7^ The sum of all assigned points was the *admission LAPS2 score*. We then similarly estimated *daily LAPS2 scores* for each ICU admission day by first building daily pre-LAPS2 models to stratify patients into high or low mortality risk daily, and then using the worst values of all LAPS2 variables from the prior ICU day to assign points to calculate the daily LAPS2 score.

### Statistical analysis

To contextualize our results within the existing evidence (i.e., performance of LAPS2 as a single score at hospital admission),^7^ we first built patient-level models using multivariable logistic regression, including hospital admission LAPS2 and covariates similar to the original specification (**Supplement**) to predict in-hospital mortality within 30 days of ICU admission. Next, we built four sets of patient-day-level models including all ICU days. First, we built logistic regression models including LAPS2 for ICU day 1 assigned to all ICU days, to simulate clinical practice where daily decisions are made with knowledge of only the admission LAPS2. Then we built logistic regression, penalized logistic regression using least absolute shrinkage and selection operator (LASSO), and random forest models that included updated daily LAPS2 for each ICU day, in order to evaluate the predictive value of adding updated daily scores and to optimize predictive ability. Hyperparameters for the penalized logistic regression using LASSO were determined by maximizing the 5-times repeated 10-fold cross validated scaled Brier score (SBS) in the training set. The SBS is a composite measure of discrimination and calibration that incorporates the difference between observed and expected outcomes and number of observations and accounts for the event rate, providing a score that is comparable across models, with higher positive values indicating better performance.^9^

For all sets of models, we used internal-external validation, such that four hospitals were included in a training set and the fifth hospital was included in a validation set, and then repeating all analyses with each hospital as the validation set. All models included the hospital admission LAPS2 and covariates similarly to the original specification of LAPS2 (**Supplement**).

We evaluated performance from fitting predicted estimates in the validation sample. The primary performance measure was the SBS. We evaluated discrimination using c-statistics. We additionally evaluated calibration by visual inspection of calibration plots.

We performed three secondary analyses. Under a hypothesis that daily LAPS2 predictive performance deteriorates over time, as physiological changes transition from acute to chronic and contribute less to short-term mortality,^10^ we evaluated the predictive performance by estimating SBS and c-statistics separately for each ICU day. To assess predictive equity, we stratified analyses by gender and race.^11^

We constructed 95% confidence intervals (CIs) using the standard deviation of estimates from bootstrapping 500 randomly selected samples. All statistical analyses were conducted using Stata version 17 (StataCorp LP, College Station, TX) and R language for statistical computing (version 4.1.2).

## RESULTS

The final study population included 13,993 patients and 120,101 ICU days. **Table 1** summarizes patient characteristics and outcomes, and **Supplementary Table 1** summarizes missingness.

**Table 1:** Characteristics and outcomes of included patients.

| Characteristic or outcome | N=13,993 |
| --- | --- |
| Age, median (IQR) | 65 (55-74) |
| Male, n (%) | 8,511 (60.8%) |
| ICU admission source, n (%) |  |
| Operating room | 6,314 (45.1%) |
| Interhospital transfer | 3,130 (22.4%) |
| Emergency department | 2,884 (20.6%) |
| Ward transfer | 1,608 (11.5%) |
| Direct admission | 57 (0.4%) |
| Hospital LOS prior to ICU admission in days, median (IQR) | 0.3 (0.1-0.9) |
| Surgical service type, n (%) | 9,268 (66.2%) |
| Elixhauser comorbidity index,<sup>3</sup> median (IQR) | 11 (4-18) |
| Mortality, n (%) | 1,657 (11.8%) |
| Hospital LOS in days, median (IQR) | 10.4 (6.09-19.8) |
| **Abbreviations:** IQR, interquartile range; ICU, intensive care unit; LOS, length of stay |  |

The patient-level model including admission LAPS2 without daily LAPS2 had an SBS of 0.175 (95% CI 0.148-0.201) and c-statistic of 0.824 (95% CI 0.808-0.840) with Hospital A as the validation set, with modest variability when validated in the other hospitals (**Supplementary Table 2**).

All models including daily LAPS2 scores performed better than the model that carried forward the single LAPS2 score from the first ICU day (with Hospital A as the validation set: SBS 0.163 [95% CI 0.151-0.174] and c-statistic 0.805 [95% CI 0.800-0.810] compared to SBS 0.096 [95% CI 0.088-0.105] and c-statistic 0.749 [95% CI 0.743-0.755]) (**Figure 1, Supplementary Table 2**). The logistic regression, penalized logistic regression using LASSO, and random forest models had similar point estimates and overlapping 95% CIs for each validation set, though performance differed among hospitals. All models were well-calibrated among patients with <50% mortality risk, representing most ICU patients (**Figure 2, Supplementary Table 2, Supplementary Figure**).

**Figure 1:**
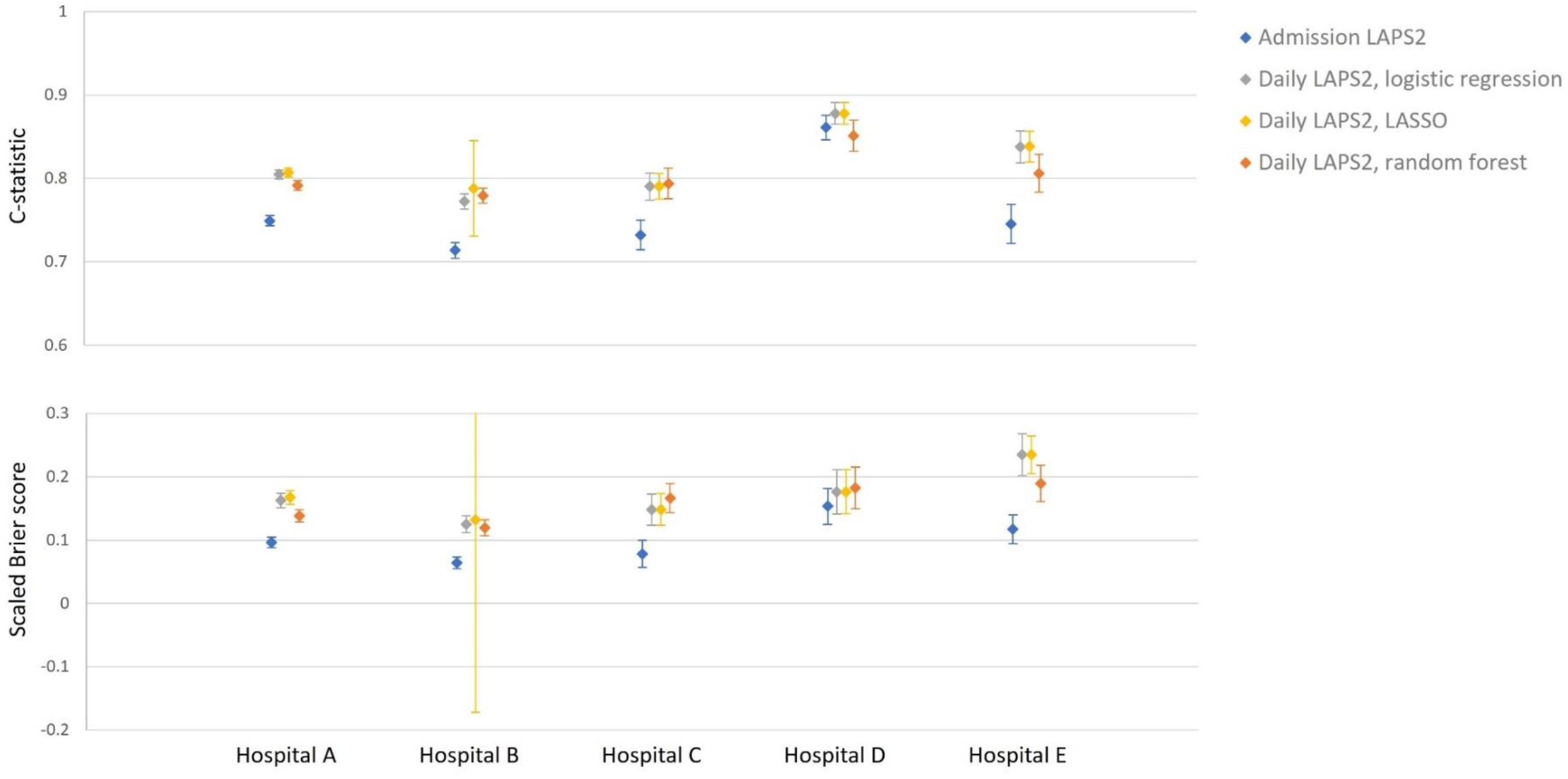
Performance of models to predict in-hospital mortality within 30 days of intensive care unit admission. **Abbreviations:** LAPS2, Laboratory-based Acute Physiology Score, version 2; LASSO, least absolute shrinkage and selection operator Closed diamonds represent point estimates for c-statistics and scaled Brier scores, as indicated. Error bars represent 95% confidence intervals, estimated by bootstrapping 500 randomly selected samples

**Figure 2:**
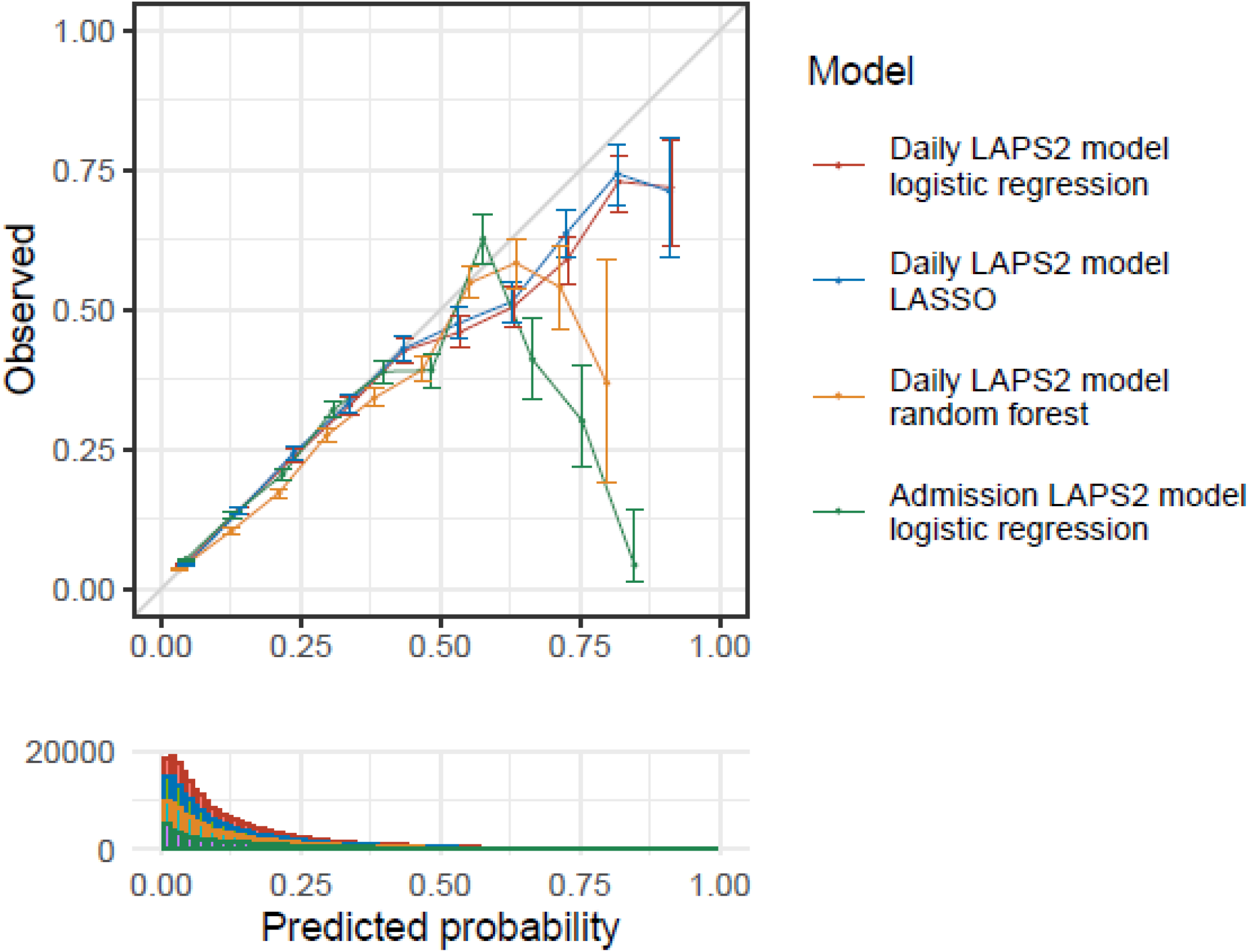
Calibration plots of models to predict in-hospital mortality within 30 days of intensive care unit admission. **Abbreviations:** LAPS2, Laboratory-based Acute Physiology Score, version 2; LASSO, least absolute shrinkage and selection operator

We performed secondary analyses using logistic regression for computational efficiency, given the findings of similar performance of the main models across modeling approaches. Models performed similarly with SBS≥0.13 and c-statistics>0.75 for most or all days through ICU day 15, with subsequent declining performance (**Figure 3, Supplementary Table 3**). Calibration differed between male and female patients in some validation sets, and between Black and White patients in most validation sets, but SBSs were not statistically different for most hospitals (**Figure 4, Supplementary Tables 4 and 5**).

**Figure 3:**
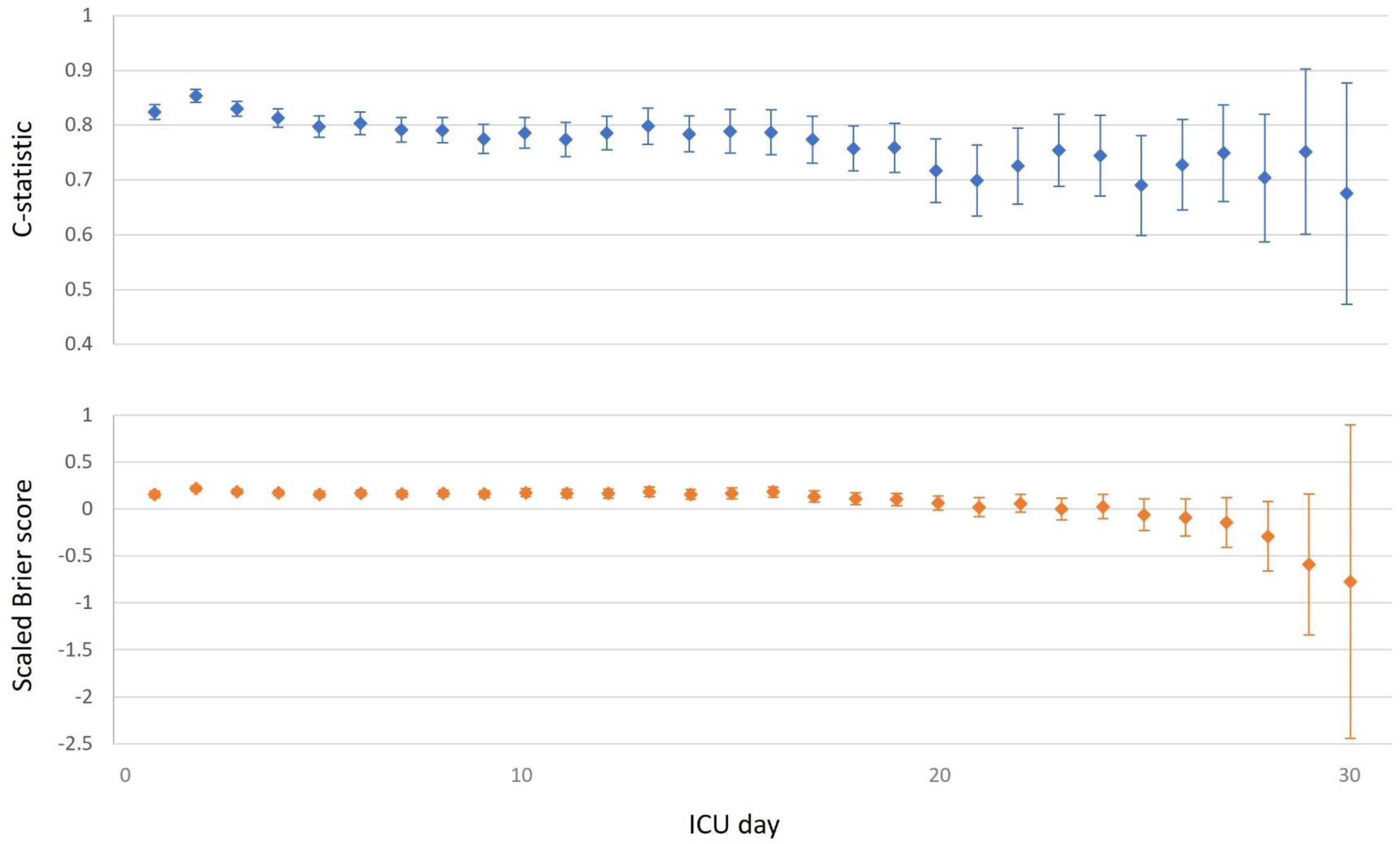
Performance of daily Laboratory-based Acute Physiology Score, version 2 (LAPS2) logistic regression model by day of intensive care unit (ICU) admission.

**Figure 4:**
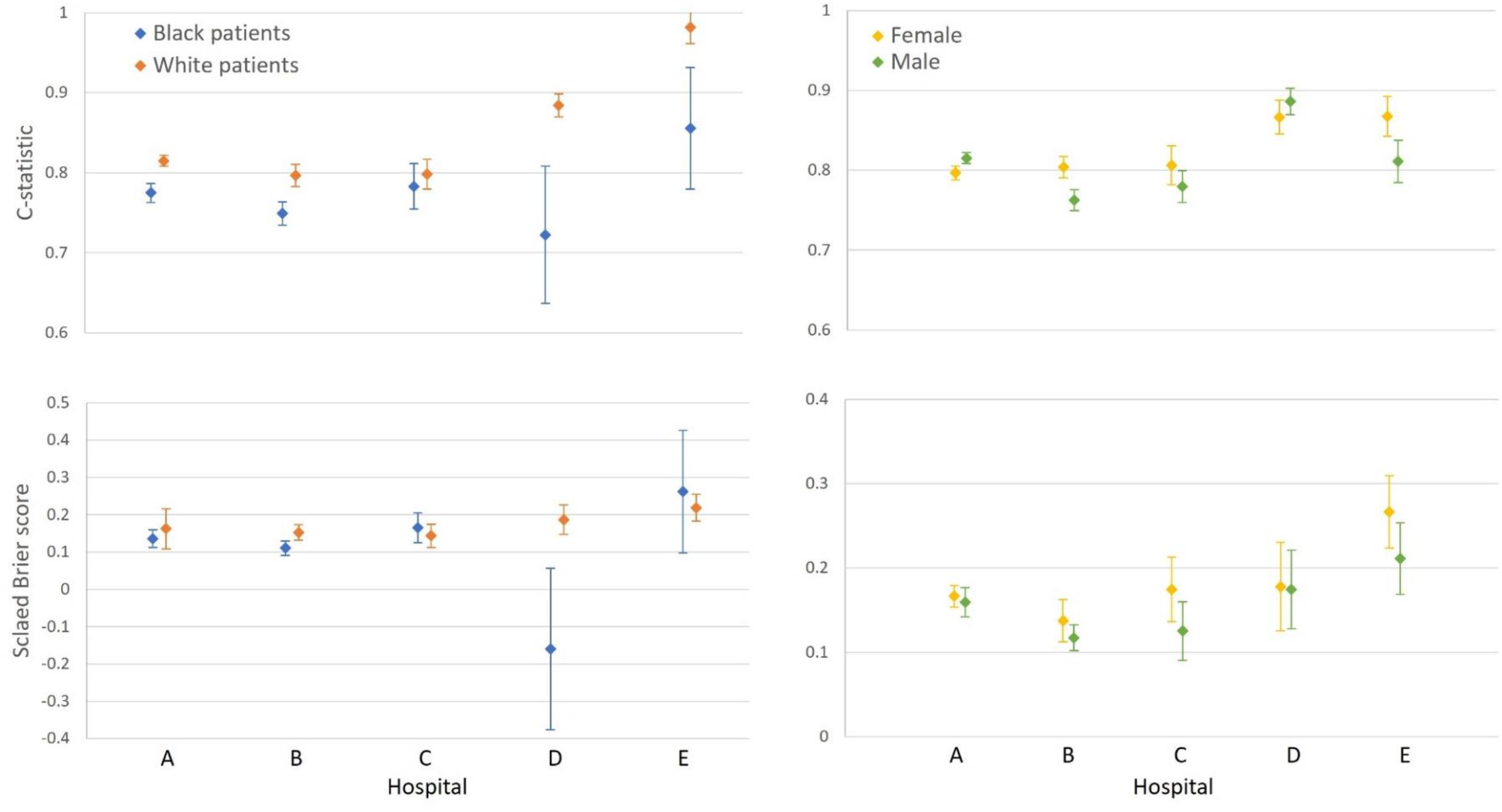
Performance of daily Laboratory-based Acute Physiology Score, version 2 (LAPS2) logistic regression model by self-reported race and gender.

## DISCUSSION

This study adds to the literature on ICU mortality prediction by adapting LAPS2 from a single hospital admission score incorporating data from the 72 hours preceding admission in an integrated health system to an admission score incorporating only data from the first 24 hours of admission plus a daily updating score. Additionally, we are not aware of prior attempts to evaluate the performance of models incorporating these LAPS2 modifications to predict in-hospital mortality among an exclusively ICU population. Our patient-level model demonstrated lower discrimination than Escobar et al.’s model.^7^ Our patient-day-level models including daily LAPS2 score improved upon this discrimination to a maximum c-statistic of 0.878. Thus, our results validate the use of modified LAPS2 scores, broadening the generalizability. Furthermore, having high-performing, time-updating mortality prediction models are critical to clinical outcome prediction, risk adjustment, quality and safety initiatives, and hospital performance.

Performance of the original LAPS2 is excellent and has been externally validated;^12–14^ however, it has several important limitations. Many health systems do not have ready access to physiologic data preceding hospital admission. Furthermore, a single score on hospital admission cannot account for physiologic changes that occur after admission and may add predictive value. Expanding the scope of LAPS2 to calculate time-updating scores to estimate mortality risk may mitigate these limitations. Indeed, one study modifying the prior version of LAPS2 by adding daily updating covariates yielded a model with excellent performance to estimate the hazard of death on each hospital day among ward patients.^15^ Another study of time-updating severity of illness scores including SOFA to predict ICU mortality similarly demonstrated high discrimination.^16^ Our results add to the evidence by demonstrating the value of daily LAPS2 scores to predict mortality on a time-updating basis among ICU patients.

While the overall performance of our time-updating LAPS2 models was strong, there was performance variability across hospitals, and calibration differed by gender and race. In particular, performance was poor among Black patients in hospitals D and E with wide CIs, likely due to data sparsity. Such variability could be problematic for clinical outcome prediction or limited resource allocation at the institutional level. Similar to Ashana et al.’s findings, overprediction of mortality among Black patients could erroneously result in deprioritization of Black patients to receive ICU beds or mechanical ventilation during mass casualty events, for example.^8^ However, other use cases such as risk adjustment or outcome assessment for ICU staffing and organizational studies may not be as adversely influenced by the race-based performance differences we observed. Future work is needed to further evaluate for algorithmic equity in similar models across health systems and to improve model equity performance.^17–24^

Additionally, while prior studies vary in “chronic critical illness” definitions from 72 hours of mechanical ventilation to 21 days of critical illness,^25^ our time-updating LAPS2 model performance highlights a decrement around ICU day 15.^10^ Although external validation is needed, we hypothesize that this may define an important transition point from acute to chronic critical illness, where persistent abnormal physiology may have lower predictive value. Having a physiologic definition of chronic critical illness could transform both clinical care and critical illness survivorship research.

Our study has several important limitations. First, we performed our study within a single health system, potentially limiting generalizability. However, our health system includes five diverse hospitals, and differs from the integrated health system where LAPS2 was originally developed. Second, we only included covariates in our prediction models that were utilized in Escobar et al.’s original manuscript for comparative purposes, but additional critical illness measures could enhance prediction. Third, LAPS2 is computationally more complex than other frequently used mortality prediction models such as SOFA. Fourth, while time-updating LAPS2 scores improved individual mortality prediction, time-updating scores may not be appropriate for other applications such as overall ICU performance. Finally, our study was not powered to assess for algorithmic equity. Larger studies are needed prior to implementing time-updating LAPS2 scores among ICU patients for clinical or research use.

## CONCLUSION

In summary, novel prediction models incorporating daily, time-updating, modified LAPS2 scores to predict mortality among an ICU population perform better than the original LAPS2. Individual daily scores used to predict 30-day in-hospital mortality perform consistently well for the first 15 days of hospitalization. Future studies should evaluate these findings in large, multicenter cohorts.

## Data Availability

All data produced in the present study are available upon reasonable request to the authors.

## Notes

**Research funding source:** Dr. Kohn is supported by NIH/NHLBI K23 HL146894. Dr. Ingraham is supported by NIH/NHLBI T32 HL07741. Dr. Anesi is supported by NIH/NHLBI K23 HL161353. Dr. Kerlin is supported by NIH/NHLBI R01 HL146386.

### Competing Interest Statement

The authors have declared no competing interest.

### Funding Statement

Dr. Kohn is supported by NIH/NHLBI K23 HL146894. Dr. Ingraham is supported by NIH/NHLBI T32 HL07741. Dr. Anesi is supported by NIH/NHLBI K23 HL161353. Dr. Kerlin is supported by NIH/NHLBI R01 HL146386.

### Author Declarations

The University of Pennsylvania Institutional Review Board gave ethical approval for this work.

