## Supplement for "Prediction of in-hospital mortality among intensive care unit patients using modified daily Laboratory-based Acute Physiology Scores, version 2 (LAPS2)"

**SUPPLEMENTARY MATERIALS**

**Table of contents**


| Section | Page |
|---|---|
| Supplementary Methods | 2 |
| Supplementary Tables | 5 |
| Supplementary Figure | 14 |

### SUPPLEMENTARY METHODS

#### Study population

All patients of age 18 years or older who were admitted to an intensive care unit (ICU) of the study hospitals during the study period were included. ICU admission was defined using the patient location in the electronic health record (EHR). For patients with multiple ICU admissions within a hospital encounter, we restricted analyses to the index admission. For patients with an inter-ICU transfer, we considered the ICU admission to include all contiguous ICU days.

#### Development of daily Laboratory-based Acute Physiology Score, version 2 (LAPS2)

*Overview*

Escobar et al. developed the Laboratory-based Acute Physiology Score (LAPS) using both administrative data and laboratory studies collected in the 24 hours preceding hospital admission to predict mortality among all hospitalized adults (including ICU patients),[2] which has been externally validated.[8] The subsequent iteration, the Laboratory-based Acute Physiology Score, version 2 (LAPS2), sought to further improve the score by (1) expanding the time horizon to 72 hours preceding hospital admission; (2) creating an additional preliminary model, the pre-LAPS2 model which subdivided the population into those with predicted mortality risks of <6% and ≥6%, thus providing a mechanism to address missing data for five score components (among patients with a predicted mortality risk <6%, missing data are imputed to normal; among patients with a predicted mortality risk ≥6%, points are assigned for missing data for arterial pH, lactate, white blood cell [WBC] count, troponin I, and neurological status); and (3) adding vital sign data to create the continuous LAPS2 score, ranging from 0 to 414.[9]

We operationalized LAPS2 variables using: arterial samples for lactate, pH, partial pressure of carbon dioxide ($PaCO_2$), and partial pressure of oxygen ($PaO_2$); serum samples for sodium (Na), total bilirubin, blood urea nitrogen (BUN), creatinine (Cr), albumin, hematocrit (Hct), WBC, anion gap (AG), bicarbonate ($HCO_3$), and troponin I (i.e., we excluded blood gas, electrophoresis, bone marrow, and pulmonary artery catheter samples); serum and point-of-care samples for glucose; and flowsheet data for temperature, heart rate (HR), respiratory rate (RR), systolic blood pressure (SBP), oxygen saturation ($O_2$ sat), and neurological score (included Glasgow Coma Scale and assessments of consciousness, mental status, orientation, pupils, Schmid Fall Risk Assessment, and speech). If a Hct value was not present at a particular timestamp but a hemoglobin value was present, the hemoglobin value was multiplied by three to estimate the Hct value. If a troponin I value was not present at a particular timestamp but a troponin T was present, troponin T was divided by ten to estimate the troponin I value. HRs greater than 250 beats per minute were excluded. RRs greater than 60 breaths per minute were excluded. Temperatures less than 85 degrees were converted to Celsius using the formula Fahrenheit*(9/5)+32, and were excluded if greater than 110 degrees Fahrenheit. We included preliminary, final, and edited final values. We excluded values that did not have a timestamp or a numerical value. Any value preceded by a greater than or less than sign was replaced with the highest or lowest category ranges from Escobar et al.'s original LAPS2 publication,[9] respectively.

*Specification of ICU day*
We considered each ICU day to start at 7:00am and end at 6:59am the following calendar day to align with clinician shift schedules. For each laboratory test and vital sign, we selected:

1. The most recent value prior to 7:00am within the prior 24 hours for variables included in the pre-LAPS2 model (Na, BUN:Cr ratio, AG:$HCO_3$ ratio) using Escobar et al.'s approach,[9] and
2. The worst value prior to 7:00am within the prior 24 hours for the LAPS2 model (minimum for pH, Na, albumin, glucose, WBC, temperature, SBP, $O_2$ sat; maximum for lactate, total bilirubin, BUN, Cr, Hct, $PaCO_2$, $PaO_2$, Troponin I, BUN:Cr ratio, HR, RR, neurological score, shock index) using Escobar et al.'s approach.[9]

*Preliminary logistic regression models (pre-LAPS2)*
All analytic models were adjusted for both hospital admission LAPS2 and ICU admission LAPS2 as hospitalization and ICU severity of illness risk adjustments, respectively. These were performed using the first 24 hours of hospital and ICU data, respectively.

In the hospital admission pre-LAPS2 model, we included a patient's age, gender, hospital admission source (emergency department [ED], direct admission, interhospital transfer), Na, BUN:Cr ratio, and AG:$HCO_3$ ratio using Escobar et al.'s approach (Escobar et al. only included ED vs other for hospital admission source).[9]

In the daily pre-LAPS2 model, we included a patient's age, gender, ICU admission source (ED, direct admission, interhospital transfer, operating room, ward), Na, BUN:Cr ratio, and AG:$HCO_3$ ratio.

For these models, if a laboratory test was missing, we imputed a normal value. When more than 1 laboratory test was available, we selected the most recent value prior to 7:00am within the prior 24 hours. For each model, categories for each variable were retained from Escobar et al.'s original publication.[9]

*LAPS2 score generation*
Once we defined the high-risk cohort for addressing missing data, we subsequently utilized the categories and associated point assignments for each variable from Escobar et al.'s original publication to generate LAPS2 scores for hospital admission, using the worst values in the 24 hours preceding 7:00am.[9] Among high-risk patients with missing data for lactate, pH, WBC, troponin I, and neurological status, points were assigned using Escobar et al.'s original publication.[9] The remaining missing data were imputed to normal, or zero points.

*Statistical analysis*
To contextualize our results within the existing evidence (i.e., LAPS2 as a single score at hospital admission), we first built patient-level models using multivariable logistic regression, including hospital admission LAPS2 and covariates similar to the original specification to predict in-hospital mortality within 30 days of ICU admission. Covariates included a patient's age, gender, ICU admission source (ED, direct admission, interhospital transfer, operating room, ward

transfer), hospital length of stay prior to ICU admission, service type (medical vs surgical), Elixhauser comorbidity index, and International Classification of Diseases, Tenth Revision admission category. Next, we built several sets of patient-day-level models including all days of ICU admission up to 30 days to compare their performance to predict in-hospital mortality within 30 days of ICU admission: (1) logistic regression including LAPS2 for ICU day 1 assigned to all ICU days, to simulate clinical practice where daily decisions are made with knowledge of only the admission LAPS2; (2) logistic regression including updated daily LAPS2 for each ICU day, to evaluate the predictive value of adding updated daily scores; and then two alternative modeling approaches to optimize the predictive ability of including daily LAPS2: (3) a least absolute shrinkage and selection operator (LASSO) model; and (4) a random forest model. Hyperparameters for the LASSO and random forest models were determined by maximizing the 5-times repeated 10-fold cross validated scaled Brier score (SBS) in the training set.

For all models, we used internal-external validation, splitting the sample such that all admissions to four hospitals were included in a training set and all admissions to the fifth hospital were included in a validation set, and then repeating all analyses for each hospital as the validation set. All models also included the hospital admission LAPS2 and covariates similar to the original specification of LAPS2 as described above.

### SUPPLEMENTARY TABLES

#### Supplementary Table 1: Missingness of Laboratory-based Acute Physiology Scores, version 2 (LAPS2) variables on intensive care unit (ICU) day 1 and for all patient-days

| Laboratory test or vital sign | ICU day 1 N=13,995 | All patient-days N=120,101 |
|---|---|---|
| pH | 2,856 (20%) | 60,038 (50%) |
| Lactate | 2,965 (21%) | 63,047 (52%) |
| Na | 122 (0.9%) | 7,791 (6%) |
| Total bilirubin | 4,888 (35%) | 66,316 (55%) |
| BUN | 134 (1%) | 7,814 (7%) |
| Cr | 129 (0.9%) | 7,803 (6%) |
| BUN/Cr | 134 (1%) | 7,817 (7%) |
| Albumin | 6,186 (44%) | 73,599 (61%) |
| Glucose | 98 (0.7%) | 4,507 (4%) |
| Hct | 128 (0.9%) | 9,091 (8%) |
| WBC | 131 (0.9%) | 9,109 (8%) |
| $PaCO_2$ | 2,851 (20%) | 60,027 (50%) |
| $PaO_2$ | 2,853 (20%) | 60,041 (50%) |
| Troponin I | 9,568 (68%) | 111,007 (92%) |
| Temperature | 239 (2%) | 1,254 (1%) |
| HR | 1,050 (8%) | 18,362 (15%) |
| RR | 25 (0.2%) | 519 (0.4%) |
| SBP | 28 (0.2%) | 170 (0.1%) |
| Shock index (HR/SBP) | 1,359 (10%) | 26,747 (22%) |
| $O_2$ sat | 32 (0.2%) | 218 (0.2%) |
| Neurological score | 1,384 (10%) | 11,797 (10%) |

**Abbreviations:** Na, sodium; BUN, blood urea nitrogen; Cr, creatinine; Hct, hematocrit; WBC, white blood cell count; $PaCO_2$, partial pressure of arterial carbon dioxide; $PaO_2$, partial pressure of arterial oxygen; HR, heart rate; RR, respiratory rate; SBP, systolic blood pressure; $O_2$ sat, oxygen saturation; AG, anion gap; $HCO_3$, bicarbonate

**Supplementary Table 2: Performance estimates for Laboratory-based Acute Physiology Scores, version 2 (LAPS2) models**

For each model, we used internal-external validation, splitting the sample such that all admission to four hospitals were included in a training set and all admissions to the fifth hospital were included in a validation set, and then repeating all analyses for each hospital as the validation set. Estimates below represent scaled Brier scores and c-statistics (95% confidence intervals) from predicted values of each hospital as the validation set, as indicated.

| | Hospital A | Hospital B | Hospital C | Hospital D | Hospital E |
|---|---|---|---|---|---|
| ***Admission LAPS2 score, logistic regression, patient-level*** | | | | | |
| Scaled Brier score | 0.175 (0.148, 0.201) | 0.136 (0.103, 0.169) | 0.109 (0.042, 0.176) | 0.161 (0.072, 0.251) | 0.141 (0.066, 0.215) |
| c-statistic | 0.824 (0.808, 0.840) | 0.777 (0.749, 0.805) | 0.774 (0.731, 0.817) | 0.867 (0.831, 0.903) | 0.768 (0.713, 0.822) |
| ***Admission LAPS2 score, logistic regression, patient-day-level*** | | | | | |
| Scaled Brier score | 0.096 (0.088, 0.105) | 0.064 (0.055, 0.073) | 0.078 (0.057, 0.100) | 0.153 (0.125, 0.182) | 0.117 (0.094, 0.140) |
| c-statistic | 0.749 (0.743, 0.755) | 0.714 (0.705, 0.723) | 0.732 (0.715, 0.750) | 0.861 (0.846, 0.876) | 0.745 (0.722, 0.769) |
| ***Daily LAPS2 score, logistic regression, patient-day-level*** | | | | | |
| Scaled Brier score | 0.163 (0.151, 0.174) | 0.125 (0.112, 0.138) | 0.148 (0.124, 0.173) | 0.176 (0.141, 0.212) | 0.235 (0.202, 0.268) |
| c-statistic | 0.805 (0.800, 0.810) | 0.772 (0.763, 0.782) | 0.790 (0.774, 0.806) | 0.878 (0.865, 0.891) | 0.838 (0.819, 0.857) |
| ***Daily LAPS2 score, least absolute shrinkage and selection operator (LASSO), patient-day-level*** | | | | | |
| Scaled Brier score | 0.167 (0.157, 0.178) | 0.132 (-0.172, 0.436) | 0.148 (0.123, 0.173) | 0.176 (0.142, 0.211) | 0.235 (0.205, 0.265) |
| c-statistic | 0.807 (0.802, 0.813) | 0.788 (0.731, 0.845) | 0.791 (0.775, 0.806) | 0.878 (0.865, 0.891) | 0.838 (0.82, 0.857) |
| ***Daily LAPS2 score, random forest, patient-day-level*** | | | | | |
| Scaled Brier score | 0.139 (0.129, 0.148) | 0.119 (0.107, 0.132) | 0.167 (0.143, 0.19) | 0.183 (0.15, 0.216) | 0.189 (0.161, 0.218) |
| c-statistic | 0.792 (0.786, 0.798) | 0.779 (0.771, 0.788) | 0.794 (0.776, 0.812) | 0.851 (0.832, 0.87) | 0.806 (0.784, 0.829) |

**Supplementary Table 3a: Performance estimates for patient-day-level logistic regression models stratified by intensive care unit (ICU) day, Hospital A**

| ICU day | N | Scaled Brier score | 95% CI | c-statistic | 95% CI |
|---|---|---|---|---|---|
| 1 | 7,808 | 0.153 | (0.119, 0.187) | 0.824 | (0.810, 0.838) |
| 2 | 7,783 | 0.219 | (0.191, 0.246) | 0.854 | (0.842, 0.866) |
| 3 | 6,138 | 0.184 | (0.158, 0.210) | 0.830 | (0.816, 0.844) |
| 4 | 4,880 | 0.170 | (0.140, 0.199) | 0.813 | (0.797, 0.830) |
| 5 | 3,952 | 0.157 | (0.125, 0.189) | 0.797 | (0.778, 0.817) |
| 6 | 3,306 | 0.167 | (0.134, 0.200) | 0.803 | (0.783, 0.824) |
| 7 | 2,840 | 0.162 | (0.126, 0.197) | 0.791 | (0.769, 0.814) |
| 8 | 2,488 | 0.167 | (0.131, 0.202) | 0.791 | (0.768, 0.814) |
| 9 | 2,197 | 0.158 | (0.120, 0.195) | 0.775 | (0.748, 0.802) |
| 10 | 1,967 | 0.174 | (0.133, 0.215) | 0.786 | (0.758, 0.814) |
| 11 | 1,746 | 0.165 | (0.118, 0.212) | 0.774 | (0.743, 0.805) |
| 12 | 1,565 | 0.164 | (0.116, 0.212) | 0.786 | (0.755, 0.817) |
| 13 | 1,400 | 0.183 | (0.131, 0.234) | 0.798 | (0.765, 0.831) |
| 14 | 1,292 | 0.157 | (0.105, 0.208) | 0.784 | (0.751, 0.817) |
| 15 | 1,188 | 0.167 | (0.110, 0.225) | 0.789 | (0.749, 0.829) |
| 16 | 1,096 | 0.181 | (0.126, 0.236) | 0.787 | (0.747, 0.827) |
| 17 | 1,007 | 0.133 | (0.072, 0.194) | 0.773 | (0.731, 0.816) |
| 18 | 938 | 0.109 | (0.046, 0.172) | 0.757 | (0.717, 0.798) |
| 19 | 877 | 0.100 | (0.033, 0.167) | 0.759 | (0.714, 0.804) |
| 20 | 811 | 0.062 | (-0.010, 0.135) | 0.717 | (0.659, 0.775) |
| 21 | 761 | 0.018 | (-0.081, 0.118) | 0.699 | (0.635, 0.764) |
| 22 | 714 | 0.059 | (-0.037, 0.156) | 0.725 | (0.656, 0.794) |
| 23 | 670 | -0.003 | (-0.118, 0.112) | 0.754 | (0.688, 0.820) |
| 24 | 625 | 0.025 | (-0.103, 0.152) | 0.744 | (0.671, 0.818) |
| 25 | 579 | -0.062 | (-0.231, 0.107) | 0.690 | (0.599, 0.781) |
| 26 | 541 | -0.089 | (-0.285, 0.106) | 0.728 | (0.645, 0.811) |
| 27 | 511 | -0.146 | (-0.409, 0.117) | 0.749 | (0.661, 0.837) |
| 28 | 466 | -0.291 | (-0.659, 0.077) | 0.704 | (0.587, 0.820) |
| 29 | 431 | -0.590 | (-1.343, 0.163) | 0.752 | (0.601, 0.902) |
| 30 | 405 | -0.775 | (-2.446, 0.896) | 0.675 | (0.473, 0.878) |

**Abbreviation:** CI, confidence interval

**Supplementary Table 3b: Performance estimates for patient-day-level logistic regression models stratified by intensive care unit (ICU) day, Hospital B**

| ICU day | N | Scaled Brier score | 95% CI | c-statistic | 95% CI |
|---|---|---|---|---|---|
| 1 | 3,877 | 0.103 | (0.053, 0.154) | 0.784 | (0.759, 0.809) |
| 2 | 3,858 | 0.205 | (0.165, 0.245) | 0.822 | (0.799, 0.845) |
| 3 | 3,095 | 0.172 | (0.132, 0.212) | 0.799 | (0.773, 0.825) |
| 4 | 2,479 | 0.143 | (0.101, 0.185) | 0.776 | (0.746, 0.805) |
| 5 | 2,039 | 0.144 | (0.100, 0.188) | 0.771 | (0.737, 0.804) |
| 6 | 1,728 | 0.122 | (0.080, 0.165) | 0.763 | (0.729, 0.796) |
| 7 | 1,498 | 0.119 | (0.068, 0.169) | 0.761 | (0.724, 0.799) |
| 8 | 1,279 | 0.122 | (0.067, 0.177) | 0.754 | (0.715, 0.794) |
| 9 | 1,129 | 0.124 | (0.060, 0.189) | 0.758 | (0.712, 0.804) |
| 10 | 1,005 | 0.143 | (0.078, 0.208) | 0.761 | (0.714, 0.809) |
| 11 | 903 | 0.106 | (0.043, 0.169) | 0.733 | (0.683, 0.784) |
| 12 | 808 | 0.077 | (0.011, 0.143) | 0.723 | (0.667, 0.779) |
| 13 | 737 | 0.076 | (0.009, 0.143) | 0.725 | (0.667, 0.782) |
| 14 | 662 | 0.070 | (-0.005, 0.145) | 0.733 | (0.672, 0.794) |
| 15 | 608 | 0.029 | (-0.045, 0.104) | 0.696 | (0.636, 0.757) |
| 16 | 557 | 0.039 | (-0.041, 0.120) | 0.721 | (0.651, 0.791) |
| 17 | 516 | 0.036 | (-0.047, 0.120) | 0.720 | (0.643, 0.796) |
| 18 | 474 | 0.059 | (-0.036, 0.154) | 0.728 | (0.651, 0.805) |
| 19 | 445 | -0.013 | (-0.099, 0.073) | 0.683 | (0.607, 0.759) |
| 20 | 410 | 0.076 | (-0.027, 0.179) | 0.738 | (0.646, 0.830) |
| 21 | 381 | 0.049 | (-0.052, 0.150) | 0.760 | (0.680, 0.840) |
| 22 | 355 | 0.052 | (-0.048, 0.153) | 0.753 | (0.666, 0.841) |
| 23 | 321 | 0.032 | (-0.103, 0.167) | 0.786 | (0.710, 0.862) |
| 24 | 298 | 0.051 | (-0.083, 0.185) | 0.761 | (0.67, 0.852) |
| 25 | 273 | -0.023 | (-0.225, 0.179) | 0.761 | (0.656, 0.866) |
| 26 | 253 | -0.057 | (-0.388, 0.273) | 0.750 | (0.645, 0.854) |
| 27 | 235 | -0.292 | (-0.973, 0.389) | 0.682 | (0.541, 0.823) |
| 28 | 217 | -0.784 | NA | 0.649 | NA |
| 29 | 199 | Negative infinity | NA | NA | NA |
| 30 | 186 | Negative infinity | NA | NA | NA |

**Abbreviations:** CI, confidence interval; NA, not applicable – estimates not available due to model non-convergence

**Supplementary Table 3c: Performance estimates for patient-day-level logistic regression models stratified by intensive care unit (ICU) day, Hospital C**

| ICU day | N | Scaled Brier score | 95% CI | c-statistic | 95% CI |
|---|---|---|---|---|---|
| 1 | 1,082 | 0.083 | (-0.020, 0.186) | 0.801 | (0.760, 0.842) |
| 2 | 1,082 | 0.196 | (0.127, 0.265) | 0.832 | (0.794, 0.871) |
| 3 | 922 | 0.185 | (0.106, 0.264) | 0.826 | (0.784, 0.868) |
| 4 | 730 | 0.185 | (0.104, 0.265) | 0.811 | (0.765, 0.857) |
| 5 | 578 | 0.137 | (0.058, 0.216) | 0.790 | (0.735, 0.845) |
| 6 | 460 | 0.176 | (0.088, 0.264) | 0.788 | (0.717, 0.858) |
| 7 | 391 | 0.165 | (0.068, 0.263) | 0.786 | (0.715, 0.858) |
| 8 | 321 | 0.160 | (0.065, 0.255) | 0.794 | (0.718, 0.870) |
| 9 | 256 | 0.129 | (-0.001, 0.259) | 0.768 | (0.676, 0.861) |
| 10 | 221 | 0.139 | (0.018, 0.260) | 0.753 | (0.653, 0.853) |
| 11 | 190 | 0.130 | (0.006, 0.254) | 0.764 | (0.667, 0.862) |
| 12 | 166 | 0.136 | (0.007, 0.265) | 0.773 | (0.665, 0.881) |
| 13 | 135 | 0.170 | (0.030, 0.309) | 0.818 | (0.722, 0.914) |
| 14 | 112 | 0.101 | (-0.057, 0.259) | 0.757 | (0.628, 0.886) |
| 15 | 94 | 0.161 | (0.004, 0.317) | 0.825 | (0.713, 0.937) |
| 16 | 76 | 0.085 | (-0.113, 0.283) | 0.711 | (0.541, 0.881) |
| 17 | 66 | 0.116 | (-0.079, 0.310) | 0.744 | (0.590, 0.898) |
| 18 | 58 | 0.110 | (-0.105, 0.324) | 0.770 | (0.626, 0.913) |
| 19 | 51 | 0.046 | (-0.161, 0.253) | 0.712 | (0.511, 0.913) |
| 20 | 39 | -0.005 | (-0.204, 0.194) | 0.758 | (0.546, 0.970) |
| 21 | 38 | 0.040 | (-0.188, 0.268) | 0.763 | (0.571, 0.954) |
| 22 | 32 | -0.053 | (-0.297, 0.191) | 0.749 | (0.561, 0.936) |
| 23 | 27 | 0.065 | NA | 0.802 | NA |
| 24 | 25 | -0.041 | NA | 0.816 | NA |
| 25 | 22 | 0.034 | NA | 0.800 | NA |
| 26 | 17 | 0.100 | NA | 0.808 | NA |
| 27 | 17 | -0.027 | NA | 0.750 | NA |
| 28 | 14 | -0.060 | NA | 0.697 | NA |
| 29 | 13 | 0.030 | NA | 0.818 | NA |
| 30 | 13 | 0.110 | NA | 0.909 | NA |

**Abbreviations:** CI, confidence interval; NA, not applicable – estimates not available due to model non-convergence

**Supplementary Table 3d: Performance estimates for patient-day level logistic regression models stratified by intensive care unit (ICU) day, Hospital D**

| ICU day | N | Scaled Brier score | 95% CI | c-statistic | 95% CI |
|---|---|---|---|---|---|
| 1 | 881 | 0.025 | (-0.140, 0.191) | 0.87 | (0.839, 0.904) |
| 2 | 874 | 0.216 | (0.131, 0.301) | 0.89 | (0.866, 0.922) |
| 3 | 789 | 0.249 | (0.177, 0.320) | 0.90 | (0.858, 0.933) |
| 4 | 684 | 0.218 | (0.138, 0.298) | 0.91 | (0.872, 0.945) |
| 5 | 571 | 0.204 | (0.110, 0.299) | 0.89 | (0.841, 0.930) |
| 6 | 448 | 0.197 | (0.090, 0.304) | 0.89 | (0.844, 0.932) |
| 7 | 321 | 0.156 | (0.028, 0.285) | 0.85 | (0.788, 0.913) |
| 8 | 229 | 0.166 | (0.045, 0.288) | 0.84 | (0.749, 0.929) |
| 9 | 176 | 0.218 | (0.089, 0.348) | 0.89 | (0.828, 0.952) |
| 10 | 133 | 0.161 | (0.030, 0.292) | 0.85 | (0.776, 0.927) |
| 11 | 108 | 0.084 | (-0.065, 0.233) | 0.79 | (0.680, 0.897) |
| 12 | 83 | 0.201 | (-0.002, 0.404) | 0.83 | (0.702, 0.967) |
| 13 | 69 | 0.013 | (-0.717, 0.743) | 0.87 | (0.766, 0.980) |
| 14 | 55 | 0.003 | NA | 0.86 | NA |
| 15 | 43 | 0.109 | NA | 0.87 | NA |
| 16 | 33 | 0.247 | NA | 0.94 | NA |
| 17 | 29 | 0.243 | NA | 0.93 | NA |
| 18 | 21 | 0.360 | NA | 0.94 | NA |
| 19 | 19 | 0.242 | NA | 0.94 | NA |
| 20 | 12 | 0.196 | NA | 0.95 | NA |
| 21 | 9 | 0.363 | NA | 1.00 | NA |
| 22 | 8 | 0.463 | NA | 1.00 | NA |
| 23 | 8 | 0.194 | NA | 1.00 | NA |
| 24 | 8 | 0.310 | NA | 1.00 | NA |
| 25 | 7 | 0.362 | NA | 1.00 | NA |
| 26 | 6 | 0.523 | NA | 1.00 | NA |
| 27 | 4 | -0.117 | NA | 1.00 | NA |
| 28 | 4 | -0.128 | NA | 1.00 | NA |
| 29 | 1 | Negative infinity | NA | NA | NA |
| 30 | 1 | Negative infinity | NA | NA | NA |

**Abbreviations:** CI, confidence interval; NA, not applicable – estimates not available due to model non-convergence

**Supplementary Table 3e: Performance estimates for patient-day level logistic regression models stratified by intensive care unit (ICU) day, Hospital E**

| ICU day | N | Scaled Brier score | 95% CI | c-statistic | 95% CI |
|---|---|---|---|---|---|
| 1 | 335 | 0.209 | (0.112, 0.306) | 0.793 | (0.74, 0.846) |
| 2 | 335 | 0.243 | (0.148, 0.338) | 0.829 | (0.777, 0.882) |
| 3 | 287 | 0.250 | (0.166, 0.335) | 0.858 | (0.806, 0.911) |
| 4 | 254 | 0.226 | (0.115, 0.337) | 0.850 | (0.792, 0.907) |
| 5 | 222 | 0.237 | (0.134, 0.340) | 0.827 | (0.750, 0.903) |
| 6 | 194 | 0.237 | (0.128, 0.345) | 0.833 | (0.76, 0.907) |
| 7 | 173 | 0.207 | (0.094, 0.320) | 0.804 | (0.706, 0.901) |
| 8 | 147 | 0.140 | (0.032, 0.249) | 0.794 | (0.706, 0.881) |
| 9 | 133 | 0.193 | (0.080, 0.306) | 0.838 | (0.747, 0.929) |
| 10 | 122 | 0.250 | (0.119, 0.381) | 0.873 | (0.800, 0.947) |
| 11 | 106 | 0.193 | (0.053, 0.334) | 0.819 | (0.693, 0.946) |
| 12 | 88 | 0.260 | (0.069, 0.451) | 0.848 | (0.744, 0.952) |
| 13 | 74 | 0.292 | (0.094, 0.491) | 0.873 | (0.774, 0.973) |
| 14 | 62 | 0.241 | NA | 0.891 | NA |
| 15 | 55 | 0.249 | NA | 0.893 | NA |
| 16 | 50 | 0.103 | NA | 0.826 | NA |
| 17 | 44 | 0.305 | NA | 0.917 | NA |
| 18 | 41 | 0.290 | NA | 0.883 | NA |
| 19 | 37 | 0.164 | NA | 0.813 | NA |
| 20 | 30 | 0.042 | NA | 0.702 | NA |
| 21 | 27 | -0.073 | NA | 0.460 | NA |
| 22 | 26 | -0.158 | NA | 0.480 | NA |
| 23 | 23 | Negative infinity | NA | NA | NA |
| 24 | 22 | Negative infinity | NA | NA | NA |
| 25 | 18 | Negative infinity | NA | NA | NA |
| 26 | 15 | Negative infinity | NA | NA | NA |
| 27 | 14 | Negative infinity | NA | NA | NA |
| 28 | 10 | Negative infinity | NA | NA | NA |
| 29 | 8 | Negative infinity | NA | NA | NA |
| 30 | 7 | Negative infinity | NA | NA | NA |

**Abbreviations:** CI, confidence interval; NA, not applicable – estimates not available due to model non-convergence

**Supplementary Table 4: Performance estimates stratified by gender for patient-day-level logistic regression model with daily Laboratory-based Acute Physiology Scores, version 2 (LAPS2), by hospital**

| Gender | N | Scaled Brier score | 95% CI | c-statistic | 95% CI |
|---|---|---|---|---|---|
| ***Hospital A*** | | | | | |
| Female | 24,866 | 0.167 | (0.154, 0.179) | 0.797 | (0.788, 0.805) |
| Male | 36,116 | 0.160 | (0.142, 0.177) | 0.815 | (0.808, 0.822) |
| ***Hospital B*** | | | | | |
| Female | 11,878 | 0.137 | (0.112, 0.163) | 0.804 | (0.791, 0.817) |
| Male | 18,947 | 0.117 | (0.102, 0.133) | 0.763 | (0.749, 0.776) |
| ***Hospital C*** | | | | | |
| Female | 3,488 | 0.175 | (0.136, 0.213) | 0.806 | (0.782, 0.831) |
| Male | 3,760 | 0.125 | (0.090, 0.160) | 0.780 | (0.760, 0.800) |
| ***Hospital D*** | | | | | |
| Female | 2,192 | 0.175 | (0.125, 0.231) | 0.867 | (0.845, 0.888) |
| Male | 3,442 | 0.125 | (0.128, 0.221) | 0.886 | (0.870, 0.903) |
| ***Hospital E*** | | | | | |
| Female | 1,294 | 0.267 | (0.224, 0.309) | 0.868 | (0.843, 0.892) |
| Male | 1,665 | 0.211 | (0.169, 0.253) | 0.811 | (0.784, 0.838) |
| **Abbreviation:** CI, confidence interval | | | | | |

**Supplementary Table 5: Performance estimates stratified by race for patient-day-level logistic regression model with daily Laboratory-based Acute Physiology Scores, version 2 (LAPS2), by hospital**

| Race | N | Scaled Brier score | 95% CI | c-statistic | 95% CI |
|---|---|---|---|---|---|
| ***Hospital A*** | | | | | |
| Black | 15,789 | 0.136 | (0.112, 0.160) | 0.775 | (0.763, 0.787) |
| White | 37,227 | 0.163 | (0.149, 0.176) | 0.815 | (0.808, 0.822) |
| ***Hospital B*** | | | | | |
| Black | 13,591 | 0.110 | (0.091, 0.130) | 0.749 | (0.734, 0.764) |
| White | 12,899 | 0.153 | (0.132, 0.173) | 0.797 | (0.783, 0.810) |
| ***Hospital C*** | | | | | |
| Black | 2,297 | 0.165 | (0.125, 0.205) | 0.783 | (0.755, 0.812) |
| White | 4,571 | 0.144 | (0.112, 0.175) | 0.798 | (0.780, 0.817) |
| ***Hospital D*** | | | | | |
| Black | 472 | -0.160 | (-0.376, 0.056) | 0.723 | (0.637, 0.808) |
| White | 4,937 | 0.187 | (0.148, 0.226) | 0.884 | (0.870, 0.898) |
| ***Hospital E*** | | | | | |
| Black | 278 | 0.262 | (0.097, 0.426) | 0.855 | (0.779, 0.931) |
| White | 2,184 | 0.218 | (0.182, 0.254) | 0.835 | (0.815, 0.855) |
| **Abbreviation:** CI, confidence interval | | | | | |

### SUPPLEMENTARY FIGURE

### Supplementary Figure: Calibration plots, hospitals B-E

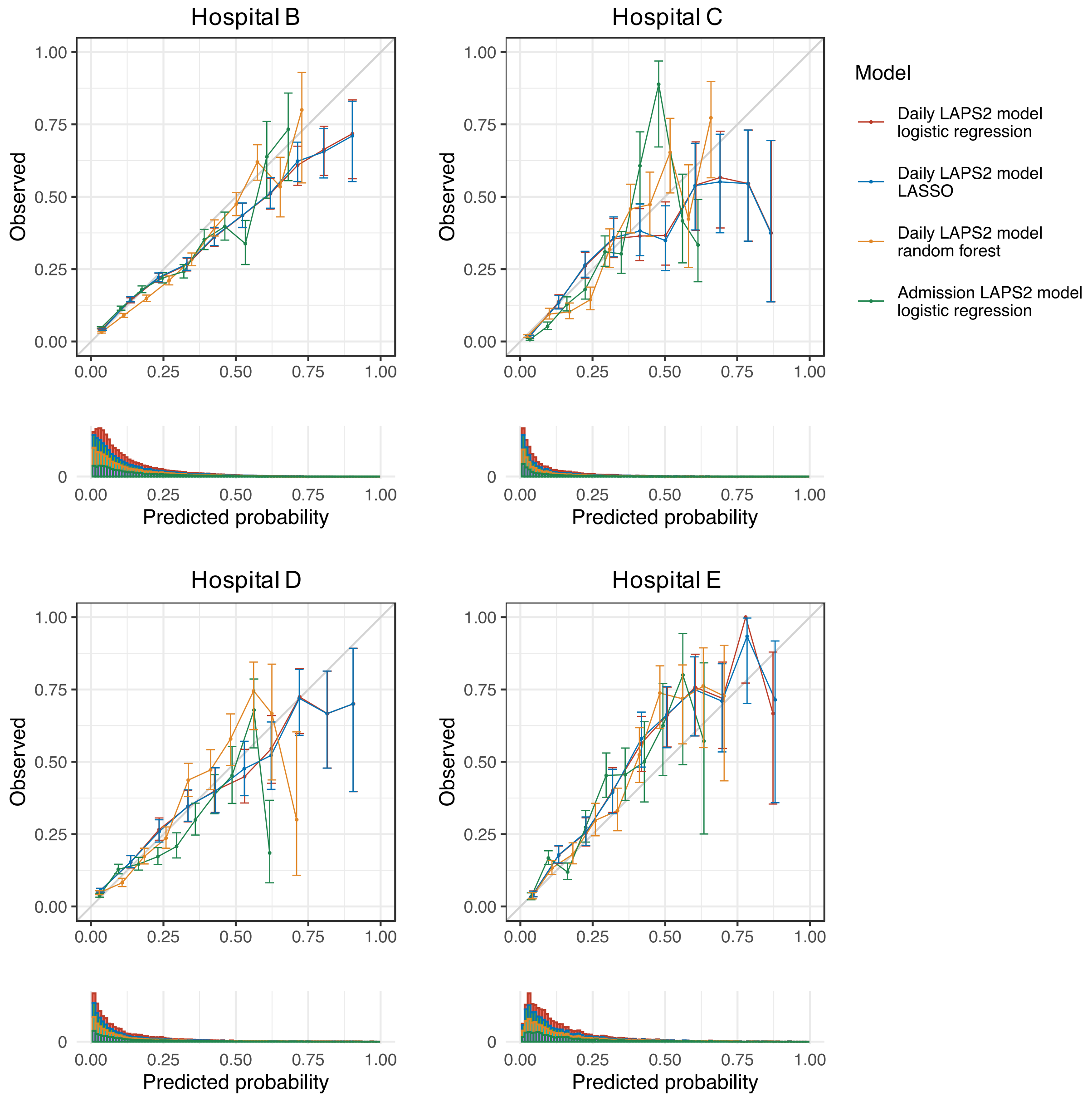
